# Application of combined genomic and transfer analyses to identify factors mediating regional spread of antibiotic resistant bacterial lineages

**DOI:** 10.1101/2020.03.03.20029447

**Authors:** Joyce Wang, Betsy Foxman, Ali Pirani, Zena Lapp, Lona Mody, Evan Snitkin

**Affiliations:** Department of Microbiology and Immunology, University of Michigan Medical School, Ann Arbor, Michigan, USA; Department of Epidemiology, University of Michigan School of Public Health, Ann Arbor, Michigan, USA; Department of Computational Medicine and Bioinformatics, University of Michigan Medical School, Ann Arbor, Michigan, USA; Division of Geriatric and Palliative Care Medicine, University of Michigan Medical School, Ann Arbor, Michigan, USA; Division of Infectious Diseases, Department of Medicine, University of Michigan Medical School, Ann Arbor, Michigan, USA

**Keywords:** Transmission, Genomic Epidemiology, Antibiotic-resistant organisms, Nursing facilities, Surveillance

## Abstract

**Background:** Patients entering nursing facilities (NFs) are frequently colonized with antibiotic resistant organisms (AROs). To understand the determinants of ARO colonization on NF admission we applied whole-genome sequencing to track the spread of four ARO species across regional NFs and evaluated patient-level characteristics and transfer acute-care hospitals (ACHs) as risk factors for colonization.

**Methods:** 584 patients from six NFs were surveyed for methicillin-resistant *Staphylococcus aureus (*MRSA), vancomycin-resistant *Enterococcus faecalis*/*faecium* (VREfc/VREfm) and ciprofloxacin-resistant *Escherichia coli* (CipREc) colonization. Genomic analysis was performed to quantify ARO spread between NFs and compared to patient-transfer networks. The association between admission colonization and patient-level variables and recent ACH exposures was examined using multivariable regression models.

**Results:** The majority of ARO isolates across study sites belonged to major healthcare-associated lineages: MRSA (ST5;N=89/117); VREfc (ST6;N=68/75); CipREc (ST131; N=58/64), and VREfm (clade A; N=129/129). While the genomic similarity of strains between NF pairs was associated with overlap in their feeder ACHs (Spearman’s rho=0.44-0.75, p<0.05 for MRSA, VREfc and CipREc), limited phylogenetic clustering by either ACH or NF supported regional endemicity. Significant predictors for ARO colonization on NF admission included lower functional status (adjusted odds ratio [aOR]>1 for all four AROs) and recent exposure to glycopeptides (aOR>2 for VREfm, VREfc and MRSA) or 3^rd^/4^th^-generation cephalosporins (aOR>2 for MRSA and VREfm). Transfer from specific ACHs was an independent risk factor for only one ARO/ACH pair (VREfm/ACH19, aOR=2.48[1.06-5.83]).

**Conclusion:** In this region, healthcare-associated ARO lineages are endemic among connected NFs and ACHs, making patient characteristics more informative of NF admission colonization risk than exposure to specific ACHs.

**Summary:** Using a combination of whole-genome sequencing, patient transfer and clinical data, we discerned the dissemination of four high-priority antibiotic-resistant organisms (ARO) in the regional healthcare network, and epidemiolocal drivers underlying the high ARO importation rate into regional nursing facilities.

## INTRODUCTION

Healthcare facilities are hotspots for the emergence and spread of antibiotic-resistant organisms (ARO) [1]. While most infection prevention efforts have focused on acute care hospitals (ACHs), the burden of ARO colonization and infection in patients in post-acute care facilities such as nursing facilities (NFs) is being increasingly recognized [2]. Since the 1990’s the use of NFs has grown dramatically to care for diverse and complex patient populations after discharge from ACHs [3,4]. NF patients are at particularly high risk of acquiring AROs because they tend to be elderly, are recovering from surgery or serious illnesses, require substantial nursing care due to the presence of medical devices and/or have limited ability to provide self-care, and are often treated with antibiotics [5]. Recent studies reported that between 30% and 65% of NF patients are colonized with at least one ARO, and 4% of NF patients are diagnosed with an ARO infection during their stay [6–8].

Like in ACHs, there is ongoing endemic transmission of AROs within NFs. In addition to within-NF transmission, NF patients are often colonized when admitted; a recent multi-site surveillance study found more than half of NF patients were colonized with at least one ARO at the time of admission [9]. This study also provided empirical evidence supporting the importance of regional dissemination of AROs via patient transfer, as nearly all patients were directly admitted from ACHs [9]. However, while a role for patient transfer is apparent, it remains unclear whether the high burden of ARO colonization entering NFs is more greatly influenced by the clinical characteristics of incoming patients or recent exposure to high-risk healthcare facilities whose infection prevention practices, antibiotic stewardship, and/or architectural design increase risk of ARO acquisition [10]. Understanding the relative contribution of these patient- and facility-level factors is essential for the design of optimal interventions to reduce ARO burden both at the level of individual healthcare facilities and across regional healthcare networks.

Here, we sought to improve our understanding of regional ARO dissemination by interrogating the distribution of ARO lineages across healthcare facilities and characterizing patient factors and prior healthcare exposures that are associated with the risk of colonization at NF admission. We focused our analysis on the four most common ARO species observed in our study NFs: methicillin-resistant *Staphylococcus aureus* (MRSA), vancomycin-resistant *Enterococcus faecalis* (VREfc), VRE *faecium* (VREfm), and ciprofloxacin-resistant *Escherichia coli* (CipREc). All four pose serious threats to public health according to the Centers for Disease Control [11]. We hypothesized that these AROs disseminate regionally with patient transfer, and that both patient clinical characteristics and recent ACH exposure contributed to the risk of colonization at the time of NF admission. By combining whole-genome sequencing (WGS), patient transfer data and clinical meta-data, we examined the relationship between ARO transmission networks and patient movement patterns, and the relative contributions of patient-level risk factors and prior healthcare exposures to the high rates of importation of a diverse set of AROs into regional NFs.

## METHODS

### Overview of Study Design

Leveraging surveillance cultures collected in a recent study [9], the current study used genomic and patient metadata to characterize the regional transmission and risk factors associated with the colonization of the four different ARO lineages simultaneously. The parent study included 651 individuals. We analyzed samples from the 584 (90% of the total) patients whose data were already complete at time of funding. For each ARO, the earliest isolate from each patient was used for whole-genome sequencing (WGS).

### Study Population

In six participating NFs in southeast Michigan, we enrolled nearly 50% of eligible patients into our study shortly after admission (mean: 5.6 days; standard deviation: 3.0 days, range: 0 -14 days). Patient age, sex, comorbidities, functional status, device use, and recent healthcare exposure were gathered at enrollment from medical notes transferred from the most recent discharge facility to NF. 95.6% of patients were directly discharged from an ACH, the remaining from another NF (3.7%), hospice (0.2%), or home (0.5%). Details about antibiotic use prior to NF admission were manually extracted from chart review, and the name of the antibiotic and start/end date of each course was recorded. We grouped 63 different antibiotics administered to enrolled patients into 27 classes. Due to low usage of most antibiotic classes, we focused our antibiotic analysis to the top three classes administered in this study population, namely 1^st^/2^nd^ generation cephalosporins (N = 61), 3^rd^/4^th^ generation cephalosporins (N = 80), and glycopeptides (N =73). Functional status was measured using the Physical Self-Maintenance Scale, ranging from full independence (score of 6) to full dependence (score of 30) in six categories of self-maintenance activities. Multiple body sites (hands, nares, oropharynx, feeding tube insertion site, suprapubic catheter site, groin, perianal area, and wounds) were cultured at enrollment, days 14 and 30, and monthly thereafter for up to 6 months. MRSA, VRE and CipREc were isolated as described [9]. All patients or his/her proxy signed a written informed consent to participate in this study, and the research was approved by the University of Michigan Institutional Review Board.

### Genomic Analysis

We extracted DNA from single colonies grown overnight in brain-heart infusion broth at 37^°^C using the PowerSoil DNA extraction kit (Qiagen). Sample-specific barcoded libraries were prepared using the Nextera DNA Flex Library Preparation Kit and sequenced using Illumina MiSeq instruments at the University of Michigan Microbial Systems Molecular Biology Laboratory. Sequences were processed to generate a final alignment file with high-confidence single-nucleotide variants (SNVs) (see Supplementary Materials for detailed methods). Sequence data are available under BioProject PRJNA435617.

### Bacterial Population Structure Analysis

We focused our analysis on dominant sequence types (STs) or genetically similar isolates that comprised the majority of the collection for each ARO species. In patients who were culture-negative at admission and became culture-positive during stay, their isolates were removed from subsequent analyses if they were closely related to another patient’s isolate from the same NF. We then used WGS data to estimate gene flow between NFs for each ARO species using an adaptation of Wright’s F statistic (Fsp) based on single nucleotide variants (SNVs) [12]. For Fsp analysis, only facilities with more than 5 isolates were included. A low Fsp suggests that two NFs share a genetically homogenous population, and *vice versa*. To evaluate genetic clustering of isolates collected from patients who were recently exposed to the same healthcare facility (ACH or NF) we carried out permutation tests to determine phylogenetic clustering by facility. See Supplementary Materials for detailed methods.

### Patient Transfer Network Analysis

At study enrolment, we first recorded the discharging hospital’s name. We then assigned a code to each hospital to ensure anonymity. The connectedness between NFs was calculated by quantifying the similarity in the distribution of transfer ACHs from where they received their patients. If two NFs had patients admitted from the same set of ACHs at identical proportions, the difference in patient transfer pattern would be zero for that NF pair. See Supplementary Materials for R packages used for calculation.

### Statistical Analysis

We used Spearman’s rank correlation to test the association between geographical distance, patient sharing-based connectedness and ARO genetic similarity. Logistic regression models were used to determine the odds ratios and associated 95% confidence intervals (95% CI) of risk factors. We constructed a separate model for each ARO and included colonized patients and patients not colonized with any ARO at enrollment as controls. We used one-way ANOVA to identify patient characteristics that differed significantly between ACHs. The final multivariate model for admission colonization was adjusted for risk factors with a p value of < 0.1 in univariate analyses. All statistical analyses were performed in R.

## RESULTS

### The burden of high-priority ARO species was due to regional dissemination of epidemic lineages

To characterize the strains circulating among regional healthcare facilities, we performed whole-genome sequencing (WGS) on the first isolate of the four high-priority AROs cultured from each colonized patient (MRSA: N=117; VREfm: N=129; VREfc: N=75; CipREc: N=64). The majority of isolates for all four ARO species were collected upon admission (**Figure S1**). Extraction of multi-locus sequencing types (MLST) from WGS data revealed that the vast majority of isolates from all six NFs belonged to sequence types (STs) commonly associated with healthcare facilities (Supplementary **Figure S2**). In particular, 76.0% of MRSA isolates belonged to ST5 and closely-related isolates (N=89/117), 90.6% of CipREc isolates belonged to a subclade of ST131, ST43, or were closely related to ST43 (N=58/64) [13], and 90.1% of VREfc isolates analyzed were ST6 (N=68/75). While multiple STs were observed for VREfm, the most dominant ST in our VREfm population, ST412, belongs to the hospital-associated clade A [14]. The close genetic relatedness of isolates in our collection suggests that the VREfm strains circulating in regional facilities were also likely members of clade A [15] (Supplementary **Figure S3**). Our subsequent analyses focused on isolates within these major healthcare-associated lineages.

### High interconnectivity among study NFs leads to a lack of genetic clustering of ARO strains by facility

The presence of common ARO lineages in all six NFs over the 3-year study period suggests these ARO strains are endemic to the regional healthcare network (**Figure 1A**). We next set out to determine if the sub-strain resolution provided by WGS would allow tracking the spread of these endemic lineages across regional facilities. To maximize our sampling of strain diversity within each NF, we included both admission isolates (MRSA: N = 67; VREfc: N = 49; VREfm: N = 101; CipREc: N = 34), and isolates from follow-up visits that were genetically distinct from any other isolate from the same NF (MRSA: N = 16; VREfc: N = 13; VREfm: N = 21; CipREc: N = 20). Examination of isolates taken from patients during follow-up visits revealed that most were not closely related to another isolate from the same NF (Supplementary **Fig S4, S5**), indicating that they were either acquired from patients not enrolled in the current study, or some patients were colonized on admission at a level below the culture detection limit. Regardless of the underlying basis, we included non-redundant isolates from admission negative patients in our subsequent analyses of regional transmission, as they provided additional information on strain diversity within each facility (See Supplementary methods for removal of redundant follow-up isolates).

**Figure 1.**
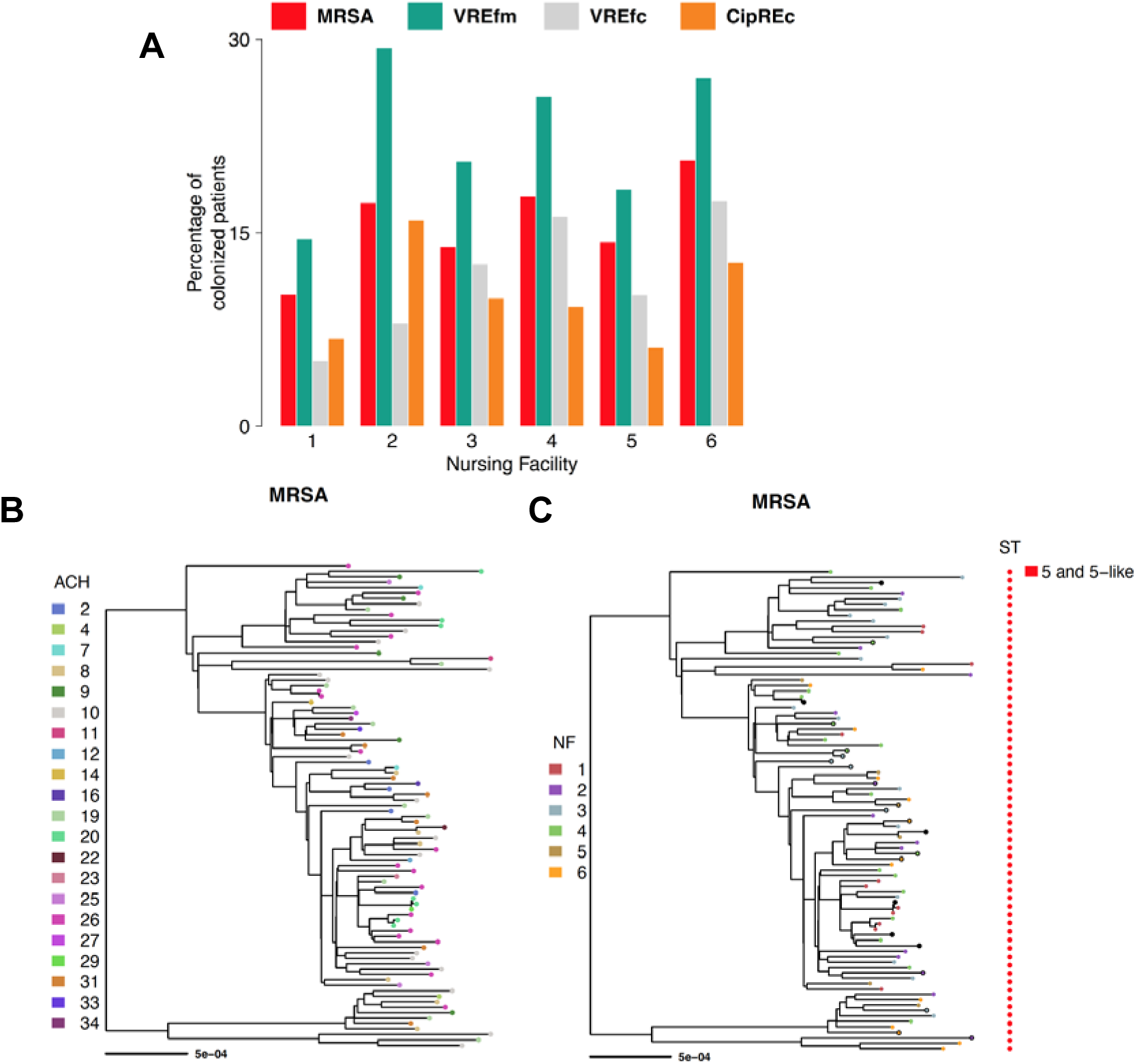
High prevalence of antibiotic resistant organisms in regional nursing facilities due to endemic spread of epidemic lineages. (A) Percentage of patients colonized with prevalent antibiotic-resistant organisms (AROs) within major healthcare lineages (MRSA: ST5 and ST5-like; VREfc: ST6; CipREc: ST43/131 and ST43/131-like, and clade A VREfm isolates) at the time of enrollment or during nursing facility stay by nursing facility. Each colour corresponds to a specific ARO or any (Total) ARO colonization prevalence. (B-C) Phylogenetic tree of MRSA isolates labeled by patient’s (B) most recent acute-care hospital (ACH) exposure and (C) patient’s nursing facility (NF) residence at the time of isolate detection. Isolates collected at the time of admission are shown as solid circles, and those collected during follow-up visits are shown as filled circles. Follow-up isolates pruned from analysis due to close genetic distance with an admission isolate within the same NF are shown as black circles. Sequence type of each isolate is shown on the right. The tree was inferred from maximum likelihood (RAxML) analysis with midpoint rooting. Scale bar represents substitutions per nucleotide site. Abbreviations: MRSA, methicillin-resistant *Staphylococcus aureus*; VREfm, vancomycin-resistant *Enterococcus faecium*; VREfc, vancomycin-resistant *Enterococcus faecalis*; CipREc, ciprofloxacin-resistant *Escherichia coli*.

We next compared genomes of ARO isolates from different NFs to attempt to track pathways of inter-facility transmission. Based on the premise that patient transfer mediates regional spread, we hypothesized that NFs receiving patients from the same feeder ACHs would harbour more genetically similar strains. However, inspection of the core-genome phylogeny did not support this hypothesis, as strains appeared highly intermixed when labeled by feeder ACHs. The lack of clustering by ACH was corroborated by phylogenetic permutation tests, which found no statistical support for clustering by most ACHs or NFs on any of the ARO core-genome phylogenies (**Figure 1B and C showing MRSA phylogeny as a representative example**, Supplementary **Figures S4, S6, S7**). To further evaluate the relationship between the genomic similarity of strains in different NFs and the overlap in their feeder ACHs, we computed the patient-sharing similarity between each pair of NFs and compared this to the genomic similarity of strains for each NF pair (**Figure 2**). While we observed a positive correlation between patient-sharing and the genomic similarity of NF strains for each ARO lineage investigated (Spearman’s rho 0.44 – 0.75, p < 0.05 for MRSA, VREfc and CipREc, p = 0.10 for VREfm), we found that the correlation was heavily driven by NF 1, which was located geographically further from the other five NFs, and had less overlap in feeder ACHs (**Figure 3**, Supplementary Figure **S8 – S10**). This suggests that while patient transfer and geographical distance influence the genetic similarity of bacterial populations between healthcare facilities, for highly interconnected facilities that are geographically proximate, the frequent intermixing of prevalent ARO lineages can preclude a clear delineation of such relationships.

**Figure 2.**
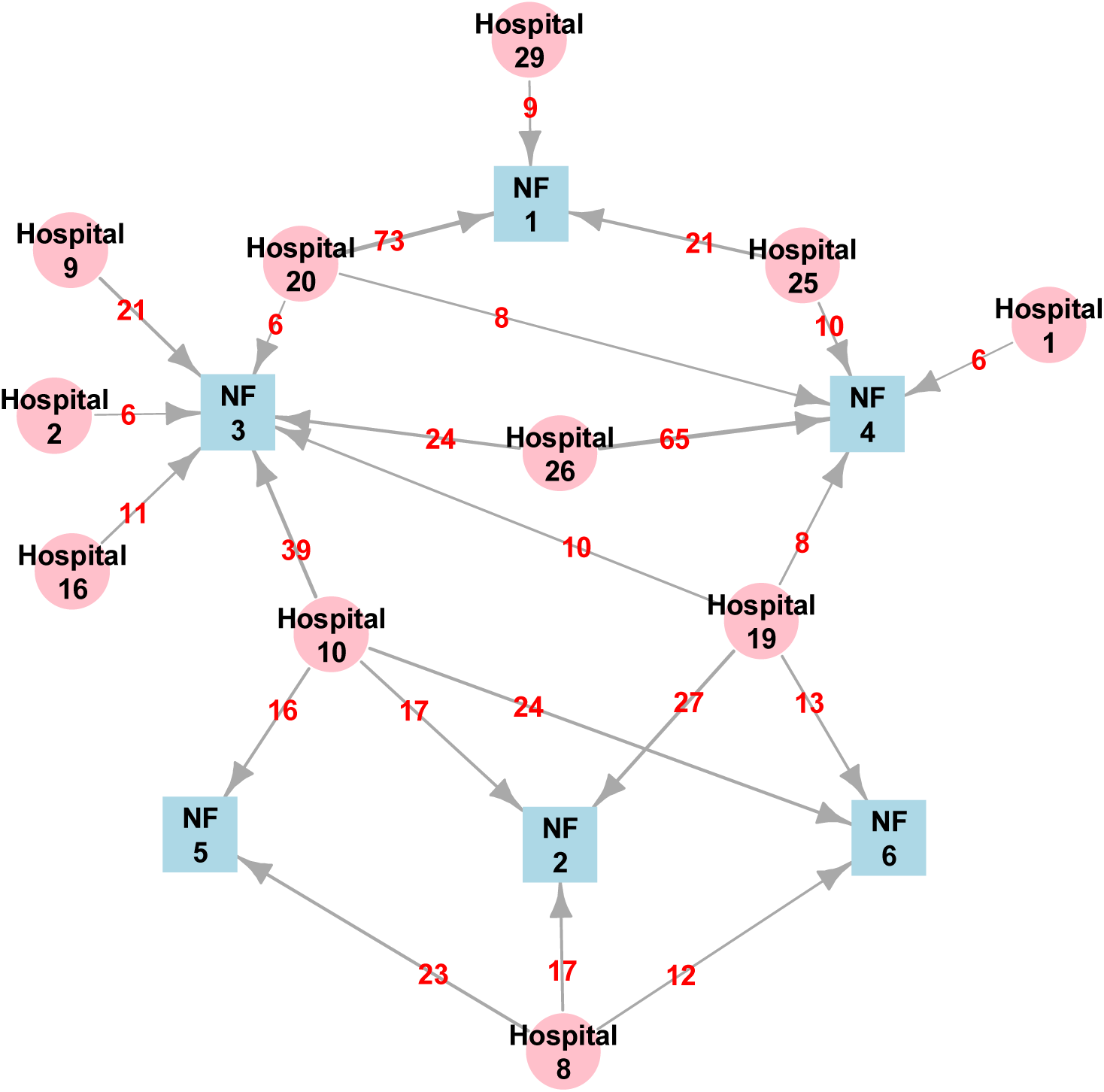
Patient sharing network between regional acute care hospitals and study nursing facilities. Visualization of patient sharing network involving six nursing facilities (NFs; blue nodes) and 11 acute-care hospitals (pink nodes) in southeast Michigan, 2013–2016. Directed arrows represent patient flow from an ACH to a NF, with the number of patients transferred shown in red. The movement of all patients enrolled in the study (N = 584) were included in this graph.

**Figure 3.**
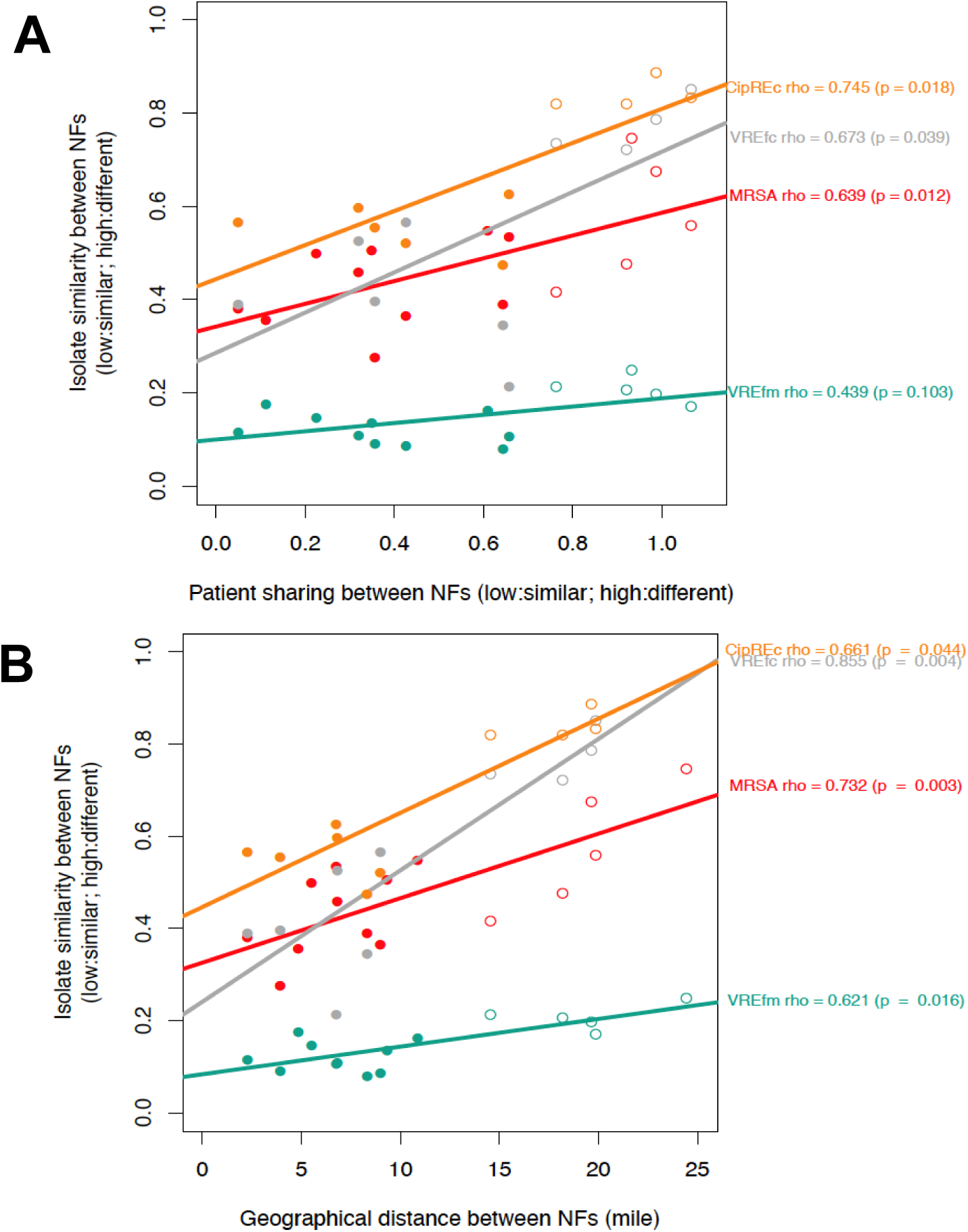
Genetic relatedness among ARO isolates from different nursing facilities associated with patient sharing and geographic proximity. Relationship between the genetic similarity of isolates between each pair of nursing facilities (NFs) and (A) overlap in feeder ACHs between the NF pair and (B) geographical distance between NF pair. Spearman’s rank correlation coefficients are shown on the right. The colour of the data points and regression lines correspond to different AROs. Closed circles denote NF pairs excluding facility 1; open circles denote NF pairs including facility 1. Abbreviations: MRSA, methicillin-resistant *Staphylococcus aureus*; VREfm, vancomycin-resistant *Enterococcus faecium*; VREfc, vancomycin-resistant *Enterococcus faecalis*; CipREc, ciprofloxacin-resistant *Escherichia coli*.

### Characteristics of patients *and* transfer hospital influence risk for ARO colonization on admission to NFs

The above results suggest that the burden of ARO colonization on NF admission for all four tested ARO species is driven by transmission of epidemic lineages at connected healthcare facilities. We next sought to understand the factors that determine which specific patients are at increased risk for ARO colonization on NF admission. In particular, we hypothesized patient risk will be influenced by a combination of the colonization pressure at connected ACHs (e.g. high-risk feeder facilities) and patients harbouring clinical features associated with risk of colonization (e.g. high-risk patients). To delineate the contribution of patient- and facility-level factors to colonization risk, we constructed an individual multivariate regression model for each ARO species, including patient characteristics with a p < 0.1 in univariate analyses and recent ACH exposure. We focused on ACHs that discharged at least 50 patients to our six study NFs collectively (ACH 8, N=55; ACH 10, N=101; ACH 19, N=61; ACH 20, N=87; ACH 26, N=90). The remaining ACHs were collapsed into an “Other” group and served as the reference facility in analyses (N = 190). Multivariate models for each individual ARO indicated a dominant role of patient factors, with lower functional status (aOR > 1 for all four AROs), exposure to glycopeptides (adjusted odds ratio; aOR > 2 for VREfm, VREfc and MRSA) and exposure to 3^rd^/4^th^-generation cephalosporins (aOR > 2 for MRSA and VREfm) being significant risk factors for colonization at admission (**Table 1**). After controlling for patient-level factors, transfer facility was a significant risk factor for only one ARO/ACH pair (VREfm/ACH19, OR = 2.48 [95% CI = 1.06-5.83]); the only other ARO/ACH pair with an OR > 2 was for CipREc/ACH10, OR = 2.27 [95% CI = 0.82 - 6.46]. However, we note that while we are treating antibiotic exposure as a patient level factor, antibiotic use was significantly different among patients transferred from different ACHs (**Table 2**), which could be indicative of antibiotic prescribing patterns at connected ACHs influencing NF admission prevalence.

**Table 1.** Associations between patient characteristics, facility exposure and colonization with an antibiotic-resistant organism (ARO) upon admission to a nursing facility by ARO. An individual multivariate regression model was built separately for each ARO. “-” indicates that the covariate was not included in the final model because the p value was ≥0.1 in the univariate analysis. Risk factors significantly associated with ARO colonization at admission are bolded. Functional status was measured by physical self-maintenance score. Hospitals with fewer than 50 discharges were collapsed and used as the referent group. Abbreviations: ACH, acute-care hospital; MRSA, methicillin-resistant *Staphylococcus aureus*; VREfm, vancomycin-resistant *Enterococcus faecium*; VREfc, vancomycin-*resistant Enterococcus faecalis*; CipREc, ciprofloxacin-resistant *Escherichia coli*, OR (95% CI) = odds ratio and 95% confidence interval.

|  | MRSA (n = 67)<br>OR (95% CI) | VREfm (n = 101)<br>OR (95% CI) | VREfc (n = 49)<br>OR (95% CI) | CipREc (n = 34)<br>OR (95% CI) |
| --- | --- | --- | --- | --- |
| Urinary catheter use in past 30 days | 1.37 (0.7 - 2.6) | 1.69 (0.94 - 3) | - | - |
| Lower functional status | **1.15 (1.08 - 1.23)** | **1.08 (1.02 - 1.15)** | **1.07 (1 - 1.14)** | **1.11 (1.02 - 1.2)** |
| Length of hospital stay | - | **1.06 (1.02 - 1.11)** | - | - |
| Charlson comorbidity score | - | 1.05 (0.91 - 1.19) | **1.17 (1.01 - 1.35)** | - |
| Exposure to 3rd/4th Gen Cephalosporins | **2.48 (1.03 - 5.81)** | **3.96 (1.97 - 8.09)** | - | - |
| Exposure to Glycopeptides | **2.95 (1.23 - 6.93)** | **2.77 (1.29 - 5.94)** | **2.62 (1.02 - 6.41)** | - |
| Hospital 8 | 0.63 (0.19 - 1.8) | 1.18 (0.45 - 2.94) | 0.43 (0.09 - 1.43) | 0.7 (0.1 - 3.03) |
| Hospital 10 | 0.77 (0.32 - 1.8) | 0.83 (0.35 - 1.9) | 0.59 (0.22 - 1.47) | 2.27 (0.82 - 6.46) |
| Hospital 19 | 1.3 (0.44 - 3.54) | **2.48 (1.06 - 5.83)** | 0.7 (0.18 - 2.18) | 1.64 (0.33 - 6.42) |
| Hospital 20 | 0.51 (0.17 - 1.35) | 0.63 (0.22 - 1.61) | 0.51 (0.16 - 1.42) | 1.53 (0.47 - 4.76) |
| Hospital 26 | 0.93 (0.37 - 2.25) | 1.6 (0.7 - 3.61) | 1.2 (0.47 - 2.92) | 1.53 (0.42 - 5.06) |

**Table 2.** A summary of the clinical characteristics of all patients discharged from each major acute-care hospital (at least 50 discharged patients) and the prevalence of antibiotic-resistant organism colonization at NF admission by discharge hospital. One-way analysis of variance (ANOVA) was used to compare patient characteristics and ARO prevalence at different hospitals.

|  | Hospital 8<br>(n = 55) | Hospital 10<br>(n = 101) | Hospital 19<br>(n = 61) | Hospital 20<br>(n = 87) | Hospital 26<br>(n = 90) | Other<br>hospitals<br>(n = 190) | P value |
| --- | --- | --- | --- | --- | --- | --- | --- |
| Male sex (%) | 45 | 43 | 44 | 44 | 36 | 42 | 0.844 |
| Urinary catheter use in<br>past 30 days (%) | 26 | 23 | 31 | 33 | 31 | 29 | 0.695 |
| Age, year (mean, SD) | 71.44<br>(13.81) | 71.71<br>(12.62) | 73.61<br>(12.96) | 77.56<br>(9.21) | 79.8 (10.46) | 74.34 (11.88) | < 0.001 |
| Physical self-maintenance scale<br>(mean, SD) | 14.45<br>(5.06) | 14.01<br>(4.41) | 13.9 (4.09) | 13.98<br>(4.13) | 15.17 (4.79) | 14.28 (4.73) | 0.467 |
| Length of hospital stay<br>(mean days, SD) | 7.84<br>(5.61) | 6.58 (5.12) | 7.9 (5.77) | 6.16 (5.3) | 6.9 (4.31) | 7.52 (9.16) | 0.478 |
| Charlson comorbidity<br>score (mean, SD) | 2.91<br>(1.89) | 3.39 (2.38) | 2.84 (1.9) | 2.13 (2.2) | 2.47 (1.97) | 2.23 (1.82) | < 0.001 |
| Black race (%) | 96 | 71 | 72 | 0 | 7 | 22 | < 0.001 |
| Exposure to 1st/2nd Gen<br>Cephalosporins (%) | 0 | 5 | 11 | 11 | 9 | 16 | 0.004 |
| Exposure to 3rd/4th Gen<br>Cephalosporins (%) | 11 | 18 | 23 | 11 | 13 | 11 | 0.145 |
| Exposure to<br>Glycopeptides (%) | 4 | 16 | 13 | 9 | 17 | 13 | 0.2 |
| MRSA colonization at<br>discharge (%) | 9 | 11 | 13 | 7 | 12 | 14 | 0.662 |

**Table 2.** A summary of the clinical characteristics of all patients discharged from each major acute-care hospital (at least 50 discharged patients) and the prevalence of antibiotic-resistant organism colonization at NF admission by discharge hospital.
|  |  |  |  |  |  |  |  |
| --- | --- | --- | --- | --- | --- | --- | --- |
| VREfm colonization at discharge (%) | 18 | 14 | 33 | 8 | 23 | 15 | 0.002 |
| VREfc colonization at discharge (%) | 5 | 8 | 7 | 6 | 11 | 10 | 0.67 |
| CipREc colonization at discharge (%) | 4 | 10 | 5 | 7 | 6 | 4 | 0.452 |

## Discussion

The lack of effective infection prevention and antibiotic stewardship programs in NFs has been hypothesized to be a major driver of the high rates of antibiotic resistance reported in NFs over the past several years [16,17]. However, an overlooked contributor to antibiotic resistance in NFs is the high rates of patients colonized with AROs entering these facilities [9,18]. Here, we focused on six regional NFs where the admission prevalence of ARO colonization was more than 50%. To gain insights into the patient populations and transmission pathways mediating the high rates of ARO colonization on admission to these NFs we integrated genomic analyses, patient transfer data and clinical information. While our data support a role for patient transfer in regional dissemination of each studied ARO species, in these regional healthcare networks individual patient characteristics - in particular physical disability and antibiotic exposure - were better predictors of ARO colonization than specific transfer facility. We hypothesize that this observation is a consequence of long-term regional endemicity of epidemic ARO lineages making the risk for ARO exposure more even across regional facilities via the constant influx of colonized patients.

MLST analysis of each ARO species revealed that most isolates found in the six regional NFs belonged to epidemic ARO lineages associated with healthcare settings [19–21]. The endemic spread of these lineages across regional networks was further demonstrated by an overall lack of phylogenetic clustering by NF or transfer ACH. We hypothesize that this lack of clustering by facility is a reflection of the high rates of inter-facility transmission leading to rapid movement of closely related strains. The rapid intermixing of strains across proximate healthcare facilities is further supported by the relative genetic isolation of ARO strains from NF 1, where the patient transfer pattern and geographical location made it most distant from the other NFs.

We observed that patient clinical characteristics were the primary risk factors for colonization with all four AROs on NF admission. Consistent with previous studies, physical disability and exposure to antibiotics were risk factors for colonization with each of the AROs studied. As antibiotic prescription patterns vary greatly across ACHs, this highlights the potential for regional antibiotic stewardship interventions to impact ARO colonization prevalence [22,23]. We note that while patient characteristics and antibiotic exposure were dominant risk factors for ARO colonization, we did observe that recent exposure to ACH 19 was an independent risk factor for VREfm colonization, suggesting that there were unmeasured facility-level factors that contributed to its association with high VREfm prevalence. This finding supports the premise that high-risk facilities in regional healthcare networks can be identified by performing admission screening at selected sentinel facilities and quantifying the independent contribution of recent facility exposures to ARO colonization risk [24]. A Chicago-based study found that transfer from long-term care hospitals was a risk factor for carbapenemase-producing *Enterobacteriaceae* colonization, motivating further investigations to identify facility practices and patient characteristics driving the risk status of different transfer hospitals [25].

We note several strengths in our study. First, the unique parent dataset allowed us to investigate the regional transmission of four different AROs simultaneously and assess the commonality of risk factors for colonization with different AROs. Second, the linkage of patient metadata to the curated patient movement data allowed us to empirically discern the risk factors for colonization unique to each organism. Importantly, curated patient transfer data allowed us to accurately assign each patient to their previous ACH, which may be difficult with aggregate data derived from billing records, especially when multiple campuses of the same hospital share the same provider identifier [26].

Our study has several limitations. First, the six NFs participating in the parent study were in metropolitan Detroit area within 25 miles of each other. The close proximity of NFs, frequent patient transfer, and a lack of data on community exposures make it challenging to delineate with certainty the location where each patient acquired ARO colonization. In particular, it is possible that the patients in our study did not acquire their ARO strains during their most recent hospitalization, but remained persistently colonized following acquisition during a prior healthcare or community exposure [27]. This possibility could account for the lack of clustering of isolates by transfer ACH. Second, we only sequenced one isolate from each patient, potentially not capturing the full genetic diversity of the bacterial population. While our approach for comparing the genetic diversity of AROs between NFs is robust to unobserved intra-patient evolution (See Methods), the failure to capture multiple independent strain acquisitions could reduce our precision.

Together, the integration of genomic and patient transfer analyses in this study provided evidence that in our regional healthcare network, regional ARO burden was driven by epidemic lineages that were endemic across the regional healthcare network. Thus, to effectively disrupt the regional transmission of AROs, healthcare facilities will need to work together to identify high-risk patients and facilities, monitor epidemiological trends and implement more effective communication strategies to control regional prevalence of these persistent resistance threats [24,28,29].

## Data Availability

Illumina reads are available on the Sequence Read Archive (SRA) in BioProject PRJNA435617.

https://www.ncbi.nlm.nih.gov/bioproject/?term=PRJNA435617

## ACKNOWLEDGMENTS

We thank members of the Mody laboratory for data collection and analysis, members of the Snitkin lab for critical discussion of genomic and epidemiological analyses, the Microbial Systems Molecular Biology Laboratory at the University of Michigan for performing whole-genome sequencing, and Drs. Rachel Slayton, Nimalie Stone, Alexander Kallen, Hannah Wolford and Paul Prabasaj (Centers for Disease Control and Prevention) on their insightful feedback on patient transfer analysis.

## REFERENCE

1. Safdar N, Maki DG. The Commonality of Risk Factors for Nosocomial Colonization and Infection with Antimicrobial-Resistant Staphylococcus aureus, Enterococcus, Gram-Negative Bacilli, Clostridium difficile, and Candida. Annals of Internal Medicine 2002; 136:834–844. Available at: https://doi.org/10.7326/0003-4819-136-11-200206040-00013. Accessed 17 July 2019.

2. Montoya A, Cassone M, Mody L. Infections in Nursing Homes. Clinics in Geriatric Medicine 2016; 32:585–607. Available at: https://linkinghub.elsevier.com/retrieve/pii/S0749069016300064. Accessed 19 November 2019.

3. Burke RE, Juarez-Colunga E, Levy C, Prochazka AV, Coleman EA, Ginde AA. Rise of Post–Acute Care Facilities as a Discharge Destination of US Hospitalizations. JAMA Intern Med 2015; 175:295–296. Available at: http://jamanetwork.com/journals/jamainternalmedicine/fullarticle/1936579. Accessed 3 August 2018.

4. Jenq GY, Tinetti ME. Post–Acute Care: Who Belongs Where? JAMA Intern Med 2015; 175:296–297. Available at: http://jamanetwork.com/journals/jamainternalmedicine/fullarticle/1936576. Accessed 24 June 2018.

5. Dumyati G, Stone ND, Nace DA, Crnich CJ, Jump RLP. Challenges and Strategies for Prevention of Multidrug- Resistant Organism Transmission in Nursing Homes. Curr Infect Dis Rep 2017; 19. Available at: https://www.ncbi.nlm.nih.gov/pmc/articles/PMC5382184/. Accessed 25 May 2019.

6. McKinnell JA, Singh RD, Miller LG, et al. The SHIELD Orange County Project: Multidrug-resistant Organism Prevalence in 21 Nursing Homes and Long-term Acute Care Facilities in Southern California. Clin Infect Dis Available at: https://academic.oup.com/cid/advance-article/doi/10.1093/cid/ciz119/5315468. Accessed 28 April 2019.

7. Mantey J, Min L, Cassone M, Gibson KE, Mody L. Changing dynamics of colonization in nursing facility patients over time: Reduction in methicillin-resistant Staphylococcus aureus (MRSA) offset by increase in vancomycin-resistant Enterococcus (VRE) prevalence. Infect Control Hosp Epidemiol 2019; :1–2.

8. Kahvecioglu D, Ramiah K, McMaughan D, et al. Multidrug-Resistant Organism Infections in US Nursing Homes: A National Study of Prevalence, Onset, and Transmission across Care Settings, October 1, 2010–December 31, 2011. Infection Control and Hospital Epidemiology 2014; 35:S48–S55. Available at: http://www.jstor.org/stable/10.1086/677835. Accessed 24 June 2018.

9. Mody L, Foxman B, Bradley S, et al. Longitudinal Assessment of Multidrug-Resistant Organisms in Newly Admitted Nursing Facility Patients: Implications for an Evolving Population. Clin Infect Dis Available at: https://academic.oup.com/cid/advance-article/doi/10.1093/cid/ciy194/4965240. Accessed 17 June 2018.

10. Drinka P, Niederman MS, El-Solh AA, Crnich CJ. Assessment of Risk Factors for Multi-Drug Resistant Organisms to Guide Empiric Antibiotic Selection in Long Term Care: A Dilemma. Journal of the American Medical Directors Association 2011; 12:321–325. Available at: http://linkinghub.elsevier.com/retrieve/pii/S152586101000215X. Accessed 2 August 2018.

11. Antibiotic Resistance Threats in the United States, 2019. 2019; :148.

12. Donker T, Reuter S, Scriberras J, et al. Population genetic structuring of methicillin-resistant Staphylococcus aureus clone EMRSA-15 within UK reflects patient referral patterns. Microbial Genomics 2017; 3. Available at: http://mgen.microbiologyresearch.org/content/journal/mgen/10.1099/mgen.0.000113. Accessed 9 May 2018.

13. Lanza VF, Toro M de, Garcillán-Barcia MP, et al. Plasmid Flux in Escherichia coli ST131 Sublineages, Analyzed by Plasmid Constellation Network (PLACNET), a New Method for Plasmid Reconstruction from Whole Genome Sequences. PLOS Genetics 2014; 10:e1004766. Available at: https://journals.plos.org/plosgenetics/article?id=10.1371/journal.pgen.1004766. Accessed 24 August 2019.

14. Raven KE, Reuter S, Reynolds R, et al. A decade of genomic history for healthcare-associated Enterococcus faecium in the United Kingdom and Ireland. Genome Res 2016; 26:1388–1396. Available at: https://www.ncbi.nlm.nih.gov/pmc/articles/PMC5052055/. Accessed 7 January 2020.

15. van Hal SJ, Ip CLC, Ansari MA, et al. Evolutionary dynamics of Enterococcus faecium reveals complex genomic relationships between isolates with independent emergence of vancomycin resistance. Microbial Genomics 2016; 2. Available at: https://www.microbiologyresearch.org/content/journal/mgen/10.1099/mgen.0.000048. Accessed 23 July 2019.

16. Mody L, Crnich C. Effects of Excessive Antibiotic Use in Nursing Homes. JAMA Intern Med 2015; 175:1339–1341. Available at: http://jamanetwork.com/journals/jamainternalmedicine/fullarticle/2337253. Accessed 26 August 2018.

17. Cohen CC, Pogorzelska-Maziarz M, Herzig CTA, et al. Infection prevention and control in nursing homes: a qualitative study of decision-making regarding isolation-based practices. BMJ Qual Saf 2015; 24:630–636. Available at: https://www.ncbi.nlm.nih.gov/pmc/articles/PMC4575834/. Accessed 23 August 2018.

18. Stone ND, Lewis DR, Lowery H, et al. Importance of bacterial burden among methicillin-resistant Staphylococcus aureus carriers in a long-term care facility. Infection control and hospital epidemiology 2008; 29:143–148.

19. Klevens RM, Morrison MA, Nadle J, et al. Invasive Methicillin-Resistant Staphylococcus aureus Infections in the United States. JAMA 2007; 298:1763–1771. Available at: https://jamanetwork.com/journals/jama/fullarticle/209197. Accessed 16 November 2019.

20. Banerjee R, Johnston B, Lohse C, Porter SB, Clabots C, Johnson JR. Escherichia coli Sequence Type 131 Is a Dominant, Antimicrobial-Resistant Clonal Group Associated with Healthcare and Elderly Hosts. Infect Control Hosp Epidemiol 2013; 34:361–369. Available at: https://www.ncbi.nlm.nih.gov/pmc/articles/PMC3916146/. Accessed 16 November 2019.

21. McBride SM, Fischetti VA, LeBlanc DJ, Jr Rcm, Gilmore MS. Genetic Diversity among Enterococcus faecalis. PLOS ONE 2007; 2:e582. Available at: https://journals.plos.org/plosone/article?id=10.1371/journal.pone.0000582. Accessed 16 November 2019.

22. MacDougall C, Polk RE. Variability in rates of use of antibacterials among 130 US hospitals and risk-adjustment models for interhospital comparison. Infect Control Hosp Epidemiol 2008; 29:203–211.

23. Polk RE, Hohmann SF, Medvedev S, Ibrahim O. Benchmarking Risk-Adjusted Adult Antibacterial Drug Use in 70 US Academic Medical Center Hospitals. Clin Infect Dis 2011; 53:1100–1110. Available at: http://academic.oup.com/cid/article/53/11/1100/306164. Accessed 26 October 2019.

24. Slayton RB, Toth D, Lee BY, et al. Vital Signs: Estimated Effects of a Coordinated Approach for Action to Reduce Antibiotic-Resistant Infections in Health Care Facilities — United States. MMWR Morb Mortal Wkly Rep 2015; 64:826–831. Available at: https://www.ncbi.nlm.nih.gov/pmc/articles/PMC4654955/. Accessed 19 July 2019.

25. Prabaker K, Lin MY, McNally M, et al. Transfer from high-acuity long-term care facilities is associated with carriage of Klebsiella pneumoniae carbapenemase-producing Enterobacteriaceae: a multihospital study. Infect Control Hosp Epidemiol 2012; 33:1193–1199.

26. Medicare C for, Baltimore MS 7500 SB, Usa M. Hospitals. 2019. Available at: https://www.cms.gov/Medicare/Provider-Enrollment-and-Certification/CertificationandComplianc/Hospitals.html. Accessed 2 August 2019.

27. Fisch J, Lansing B, Wang L, et al. New Acquisition of Antibiotic-Resistant Organisms in Skilled Nursing Facilities. J Clin Microbiol 2012; 50:1698–1703. Available at: http://jcm.asm.org/content/50/5/1698. Accessed 10 December 2014.

28. Evans CT, Jump RL, Krein SL, et al. Setting a Research Agenda in Prevention of Healthcare-Associated Infections (HAIs) and Multidrug-Resistant Organisms (MDROs) Outside of Acute Care Settings. Infection Control & Hospital Epidemiology 2018; 39:210–213. Available at: http://www.cambridge.org/core/journals/infection-control-and-hospital-epidemiology/article/setting-a-research-agenda-in-prevention-of-healthcareassociated-infections-hais-and-multidrugresistant-organisms-mdros-outside-of-acute-care-settings/3F6FB463929EC2002A115B100C2CECE1/corereader. Accessed 25 August 2018.

29. Mody L, Washer L, Flanders S. Can Infection Prevention Programs in Hospitals and Nursing Facilities Be Integrated?: From Silos to Partners. JAMA 2018; 319:1089–1090. Available at: http://jamanetwork.com/journals/jama/fullarticle/2674119. Accessed 3 August 2018.

